# Neighborhood Disadvantage, Identity-based Resilience, and Hypertension: The Healthy Aging with Resilient Identities (HARI) Study

**DOI:** 10.64898/2026.09.17.26363316

**Authors:** Hannah Pleasants, Ganga S. Bey

## Abstract

**Introduction:** Perceived neighborhood problems (PNP), such as safety concerns and limited resources, have been linked to elevated hypertension risk through chronic stress pathways. The Identity Vitality Pathology (IVP) framework captures the extent to which individuals experience threats to identity, belonging, and psychosocial vitality in the face of structural disadvantage, and may modify these pathways.

**Methods:** We conducted logistic regression analyses to estimate the association between PNP and hypertension. Models were stratified by race and gender groups (Black women, Black men, White women, White men). Both main effects models and adjusted models included an interaction term between PNP and Identity Vitality Pathology score (IVPS) to assess effect modification. Adjusted models controlled for age, education, perceived support, financial strain, physical activity, diabetes, smoking status, and self-rated health. Interaction terms were used to evaluate whether the association between PNP and hypertension varied by levels of IVPS within each race/gender group.

**Results:** Higher PNP was associated with increased odds of hypertension among White women and men, but not consistently among Black women and men. Identity state moderated this association only among Black women, such that each 1-SD increase in IVPS score was associated with 59% lower odds of hypertension at the highest level of PNP (aOR-0.41; 95% CI: 0.19, 0.88), with no meaningful moderation observed in other groups.

**Conclusion:** Identity state may moderate the association between subjective neighborhood disadvantage and hypertension, particularly among Black women, representing a potentially novel, modifiable protective factor among multiply marginalized persons living in communities with increased risk for hypertension.

## Introduction

Hypertension burdens 46.7% of the United States, with stark disparities at the intersections of ethnicity, race, and gender [1]. In 2020, nationally representative data showed that non-Hispanic (NH) White women and men had hypertension prevalences of 40% and 47%, respectively, while approximately 56% of NH Black women and men had hypertension [1]. Further, although blood pressure values trended downward from 1999 to 2010 and then upward again from 2010 to 2020 across most intersectional groups, NH Black women were the only group in which the prevalence of hypertension was higher in 2020 than in 1999 [1]. Targeted public health intervention facilitated improvement in blood pressure control among NH Black adults from 2017 to 2023 [2], however, disparities persist within hypertension-related mortality, with NH Black adults having over twice the odds as NH White adults of hypertensive in-hospital mortality [3]. These disparities are not fully accounted for by traditional cardiovascular risk factors alone, as evidence points to distinct pathways through which the structural and environmental conditions in which people live shape hypertension risk across race and gender [4,5].

Racial residential segregation and a long history of racialized disinvestment have systematically concentrated Black Americans into structurally disadvantaged neighborhoods [6,7], shaping the conditions under which health within these populations is produced and maintained across the life course [8,9]. These marginalized built environments are characterized by reduced access to quality food [10], healthcare [11], and spaces for physical activity [12], and disproportionately targeted by nicotine and alcohol marketing, key influences on cardiovascular health [13–15].

Neighborhood socioeconomic status has been shown to explain roughly 19% of the association between racial residential segregation and hypertension [4], and neighborhood characteristics including walkability, social cohesion, food environment quality, and safety have each independently been associated with hypertension prevalence [4,16]. While these objective measures identify important neighborhood characteristics, they do not capture residents’ day-to-day experiences of these conditions that may directly influence physiological stress responses that have been linked to blood pressure by a wealth of evidence [16,17]. Perceptions of safety, crime, physical disorder, and social cohesion capture a experiential domain of disadvantage that objectively-measured neighborhood features cannot capture. Because physiological stress responses are initiated through cognitive appraisal of threat rather than objective exposure to stressors alone [18], perceptions of disadvantage could act as a more proximal determinant of hypertension risk than objective socioeconomic indices, though this has not been directly tested. Consistent with this hypothesis, perceived neighborhood conditions have shown stronger associations with depressive symptoms, and the downstream inflammatory markers relevant to cardiovascular risk, than objective neighborhood characteristics [19].

The pathways through which perceptions of neighborhood disadvantage become embodied as hypertension likely involve interconnected physiological, behavioral, and structural mechanisms. Chronic exposure to neighborhood stressors may activate the hypothalamic-pituitary-adrenal (HPA) axis and sympathetic nervous system, resulting in sustained elevations of stress hormones [20]. Elevated levels of norepinephrine, epinephrine, dopamine, and cortisol have each been associated with increased risk of incident hypertension [21], and may, in turn, reflect the physiological consequences of persistent exposure to neighborhood-level stressors that dysregulate blood pressure over time. Within this framework, allostasis refers to the processes through which physiological systems maintain stability in response to environmental change and stress [22], whereas allostatic load reflects the cumulative physiological burden of repeated or chronic stress activation [23]. Persistent disruption of allostasis may therefore contribute to hypertension through prolonged neuroendocrine and autonomic dysregulation. Chronic stress may also operate through behavioral pathways, prompting coping responses that independently increase cardiovascular risk. For example, consumption of carbohydrate– and fat-rich foods may be neurobiologically reinforced by attenuating HPA axis activation [24,25], while nicotine use can increase circulating stress hormones [26]. Importantly, these behavioral responses are shaped and reinforced by the broader structural environment: marginalized neighborhoods may provide fewer opportunities for health-promoting behaviors while simultaneously increasing exposure to tobacco and alcohol marketing [13–15]. Thus, the embodiment of neighborhood disadvantage may arise through mutually reinforcing physiological and behavioral pathways, collectively contributing to the elevated burden of hypertension observed among Black Americans [27–30].

Modifiable protective factors have been found to reduce the risk of cardiovascular outcomes even in the context of neighborhood disadvantage [31,32], suggesting that there are accessible resources that may attenuate the effects of structurally-driven neighborhood stressors and cardiovascular health. For instance, prior work from our group found that optimism reduces the risk of incident cardiovascular disease specifically in the context of neighborhood disadvantage. When further stratified by ethnoracial group, this protective association remained only among Black participants, underscoring the importance of identifying the unique factors underpinning psychosocial resilience in marginalized communities [33]. Additional protective factors have been found to buffer the impact of neighborhood disadvantage on hypertension, namely whether persons perceive the positives of their neighborhood as outweighing the adversities [34] and whether persons recently volunteered at a local social organization [35]. However, existing resilience frameworks have not fully articulated the mechanisms by which psychosocial resilience can modify the perception of, and physiological response to, chronic social stressors.

The Identity Vitality Pathology (IVP) framework addresses this gap by conceptualizing *identity state* as a modifiable psychosocial construct that moderates one’s stress perception and subsequent physiological response [36,37]. Within this framework on end of the identity state spectrum is a vitalized identity, characterized by an inclusive self-concept, the belief in an intrinsic basis of value for all living beings, and unconditional compassion for all living beings including the self that stems fundamentally from a disidentification with the physical body as the predominant self. Identity vitality is theorized to attenuate the impact of chronic stressors on adverse health outcomes, including hypertension, by influencing how indviduals appraise and respond to stressors. Conversely, on the other end of the identity state spectrum, a pathologized identity state may amplify stress reactivity and reduce capacity for adaptive coping, thereby intensifying the physiological consequences of chronic adversity. Distinct from individual positive psychological traits, identity state is posited as an identity-based orientation hypothesized as an upstream resilience phenotype that influences an array of adaptive psychological characteristics that in turn shape cognitive, emotional, and behavioral responses to stress.

The IVP framework proposes that identity-based resilience stemming from identity vitality may be particularly impactful in mitigating the specific manifestations of chronic stressor exposure that are unique to specific intersectional populations [37]. For example, a compelling body of literature has identified unique cardiometabolic responses to stress among Black women, theorized as partially attributable to group-specific stressor exposure[38,39] and culturally influenced coping behavior [40], including the consumption of food high in fat and high processed sugar as well as greater reliance on social support [41,42]. Thus, IVP theory positions identity vitality as likely to have a greater protective influence on hypertension risk among Black women than other groups located at the intersections of race and gender.

Despite a growing literature linking neighborhood context to cardiovascular outcomes, few studies have examined how resident perception of neighborhood problems is associated with hypertension, and none have considered identity state as a moderator of this association. Using data from the Healthy Aging with Resilient Identities (HARI) study, we examined the association between perceived neighborhood problems and prevalent hypertension, and evaluated whether identity state moderated this association. Given documented heterogeneity across intersectional identities in both neighborhood exposures and psychosocial resilience resources [33,43], analyses were stratified by race-gender group to assess whether the moderating role of identity state differed across intersecting social positions.

## Methods

### Study population

Data for this analysis were drawn from the HARI study, a National Institute on Aging-funded cross-sectional study conducted in 2025. A nationally distributed sample of 2,011 adults was recruited over a one-month period via online survey in partnership with a health research participant recruiting firm. Participants were self-identified non-Hispanic Black and white women and men between the ages of 35 and 65 years, recruited across all four U.S. Census regions (Northeast, South, Midwest, and West) with representation from every state. All participants provided electronic confirmation of informed consent prior to completing the survey. The study protocol was approved by the University of North Carolina Institutional Review Board. Three participants who identified as non-binary gender were excluded from the present analysis, yielding a final analytic sample of 2,008. No additional exclusions were applied. The analytic sample was stratified by race and gender, resulting in four groups: Black women (n=500), Black men (n=502), white women (n=503), and white men (n=503).

### Outcome: Hypertension

The primary outcome was hypertension assessed via self-report. Participants were asked whether they had ever been told by a medical professional that they have high blood pressure or hypertension. Responses were coded as a binary variable (yes/no).

### Exposure: Perceived Neighborhood Problems

Subjective neighborhood disadvantage was assessed using a single item adapted from the validated Perceived Neighborhood Problems (PNP) scale [44], capturing perceptions of noise, traffic, speeding, and the persistence of trash and litter. Participants rated their agreement with the statement “my neighborhood has a serious problem with one or more of the following issues: excessive noise, heavy traffic, speeding cars, trash and/or litter” on a 4-point Likert scale ranging from strongly agree to strongly disagree. For regression analyses, PNP was operationalized as a four-level categorical variable—none (strongly disagree), low (disagree), moderate (agree), and high (strongly agree)—with none as the referent category.

### Moderator: Identity State

Identity state was assessed using the validated 20-item Identity Vitality Pathology Scale (IVPS-20) [36]. The IVPS measures placement along the theorized multidimensional spectrum from identity pathology to identity vitality (**Figure 1**) [36]. Identity vitality is characterized by a predominant self-concept that is inclusive of all living beings, a belief in the inherent worth of all living beings, and unconditional compassion for all living beings, including the self. Participants rated each item on a 4-point Likert scale ranging from strongly agree to strongly disagree on each item. Example items include “what people think of me doesn’t much impact who I am,” “though we may look different on the outside, at the core I am the same as everyone else,” and “even people I don’t know deserve my concern.” Item ratings were summed to produce a continuous composite score with higher scores reflecting greater identity vitality. For the present analysis, IVPS was modeled as a continuous variable standardized to a mean of 0 and a standard deviation of 1.

**Fig. 1.**
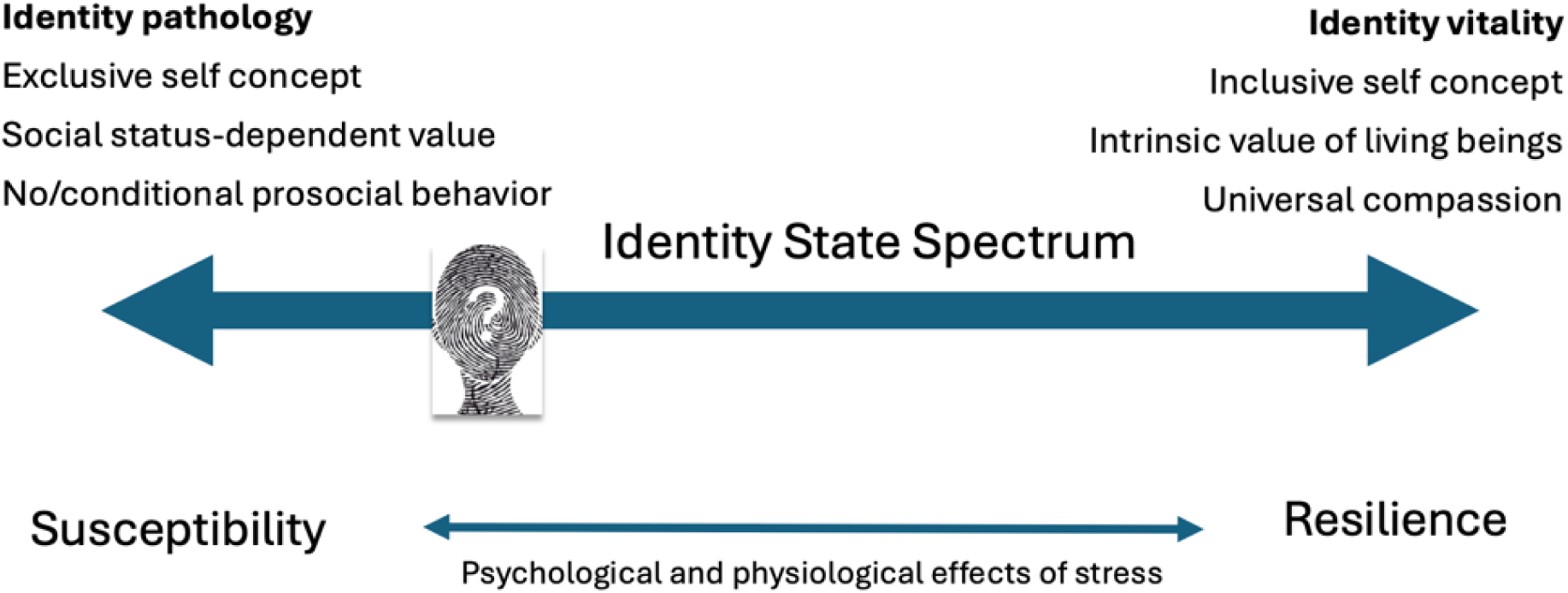
The multidimensional Identity State spectrum as described by Identity Vitality-Pathology theory spans identity pathology to identity vitality. The three dimensions of the identity state spectrum encompass exclusive self-concept vs. inclusive of all living beings; perceived status-dependent vs. intrinsic value of living beings (including the self); and propensity toward minimizing suffering of all living beings. Greater identity vitality is posited as a biologically embedded but modifiable resilience phenotype that mitigates the adverse physiological and psychological effects of chronic stress

### Covariates

Age was included as a continuous variable based on participant self-report. Individual socioeconomic status was ascertained using educational attainment and perceived chronic financial strain. Educational attainment was self-reported as highest degree earned and collapsed into five categories for analysis: less than or equal to high school, some college or technical schooling, two-year college degree, four-year college degree, and graduate degree (master’s, doctoral, or equivalent). Percived chronic financial strain was assessed with a single item asking, “on a regular basis, how well are you able to meet your basic financial needs?” with four response options ranging from no difficulty to regularly struggling. Perceived financial support was assessed using a 7-item Likert scale measuring agreement with the statement “if I run into financial difficulties, I can rely on others to support me,” reflecting the degree to which participants perceived peer-based financial support to be available to them.

Health behaviors were assessed as smoking behavior and physical activity. Smoking status was self-reported and categorized as never, former, or current smoker. Physical activity was derived from the International Physical Activity Questionnaire (IPAQ) [45]. Weekly minutes of moderate– and vigorous-intensity activity were calculated by multiplying reported days of activity by average minutes per session. Participants were then classified using a binary variable indicating whether they met CDC guidelines for physical activity, which are defined as at least 150 minutes per week of moderate-intensity aerobic activity or at least 75 minutes per week of vigorous-intensity aerobic activity [46].

Health status covariates included self-reported diabetes and self-rated health. Diabetes was operationalized as a binary variable (yes/no) based on whether a participant had ever been told by a medical professional that they have diabetes. Self-rated health was assessed with the single item “in general, would you say that your health is excellent, very good, good, fair, or poor?” and retained as an ordinal variable in analyses.

### Statistical Analysis

The association between perceived neighborhood problems and hypertension was examined separately within each of the four race-gender groups using logistic regression models. For each group, three sequential model specifications were estimated. Model 1 was unadjusted. Model 2 adjusted for sociodemographic and socioeconomic factors including age, educational attainment, perceived financial support, and financial strain. Model 3 further adjusted for health behaviors and health status indicators including smoking status, diabetes, self-rated health, and physical activity.

To evaluate whether identity state moderated the association between PNP and hypertension, each of the three model specifications were re-estimated with the addition of interaction terms between each level of PNP and the standardized IVPS score (z-score). Interaction was assessed on the multiplicative scale, and the precision of confidence intervals around interaction terms was used to evaluate the consistency and direction of effect modification. Interaction odds ratios reflect the change in odds ratio for the PNP-hypertension association per one-standard-deviation increase in IVPS. All analyses were conducted separately by race-gender group to allow for heterogeneity in both main effects and interaction patterns across groups. All analyses were conducted in SAS version 9.4 (SAS Institute, Cary, NC).

## Results

### Descriptive Statistics

Table 1 presents baseline characteristics stratified by race-gender groups. Across all groups, mean age ranged from 46.4 (SD=8.3) in Black men to 50.7 (SD=9.5) in White women. IVPS scores were highest among Black women (mean=48.1; SD=8.3) and lowest among White men (mean=43.9; SD=9.1).

**Table 1:**
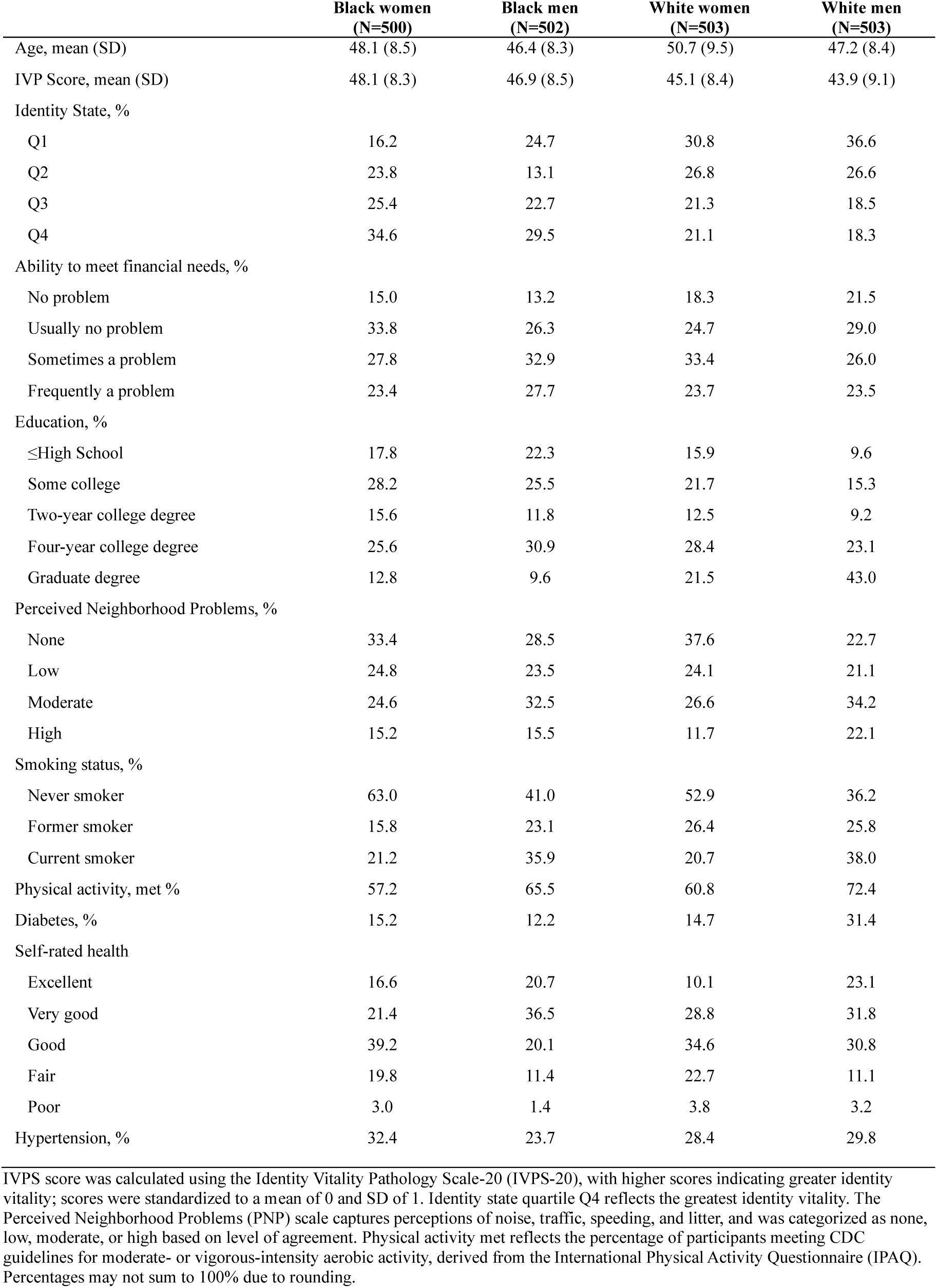
Descriptive Statistics of Participants in the Healthy Aging and Resilient Identities Cohort (N=2,008)

|  | **Black women**<br>**(N=500)** | **Black men**<br>**(N=502)** | **White women**<br>**(N=503)** | **White men**<br>**(N=503)** |
| --- | --- | --- | --- | --- |
| Age, mean (SD) | 48.1 (8.5) | 46.4 (8.3) | 50.7 (9.5) | 47.2 (8.4) |
| IVP Score, mean (SD) | 48.1 (8.3) | 46.9 (8.5) | 45.1 (8.4) | 43.9 (9.1) |
| Identity State, % |  |  |  |  |
| Q1 | 16.2 | 24.7 | 30.8 | 36.6 |
| Q2 | 23.8 | 13.1 | 26.8 | 26.6 |
| Q3 | 25.4 | 22.7 | 21.3 | 18.5 |
| Q4 | 34.6 | 29.5 | 21.1 | 18.3 |
| Ability to meet financial needs, % |  |  |  |  |
| No problem | 15.0 | 13.2 | 18.3 | 21.5 |
| Usually no problem | 33.8 | 26.3 | 24.7 | 29.0 |
| Sometimes a problem | 27.8 | 32.9 | 33.4 | 26.0 |
| Frequently a problem | 23.4 | 27.7 | 23.7 | 23.5 |
| Education, % |  |  |  |  |
| ≤High School | 17.8 | 22.3 | 15.9 | 9.6 |
| Some college | 28.2 | 25.5 | 21.7 | 15.3 |
| Two-year college degree | 15.6 | 11.8 | 12.5 | 9.2 |
| Four-year college degree | 25.6 | 30.9 | 28.4 | 23.1 |
| Graduate degree | 12.8 | 9.6 | 21.5 | 43.0 |
| Perceived Neighborhood Problems, % |  |  |  |  |
| None | 33.4 | 28.5 | 37.6 | 22.7 |
| Low | 24.8 | 23.5 | 24.1 | 21.1 |
| Moderate | 24.6 | 32.5 | 26.6 | 34.2 |
| High | 15.2 | 15.5 | 11.7 | 22.1 |
| Smoking status, % |  |  |  |  |
| Never smoker | 63.0 | 41.0 | 52.9 | 36.2 |
| Former smoker | 15.8 | 23.1 | 26.4 | 25.8 |
| Current smoker | 21.2 | 35.9 | 20.7 | 38.0 |
| Physical activity, met % | 57.2 | 65.5 | 60.8 | 72.4 |
| Diabetes, % | 15.2 | 12.2 | 14.7 | 31.4 |
| Self-rated health |  |  |  |  |
| Excellent | 16.6 | 20.7 | 10.1 | 23.1 |
| Very good | 21.4 | 36.5 | 28.8 | 31.8 |
| Good | 39.2 | 20.1 | 34.6 | 30.8 |
| Fair | 19.8 | 11.4 | 22.7 | 11.1 |
| Poor | 3.0 | 1.4 | 3.8 | 3.2 |
| Hypertension, % | 32.4 | 23.7 | 28.4 | 29.8 |

IVPS score was calculated using the Identity Vitality Pathology Scale-20 (IVPS-20), with higher scores indicating greater identity vitality; scores were standardized to a mean of 0 and SD of 1. Identity state quartile Q4 reflects the greatest identity vitality. The Perceived Neighborhood Problems (PNP) scale captures perceptions of noise, traffic, speeding, and litter, and was categorized as none, low, moderate, or high based on level of agreement. Physical activity met reflects the percentage of participants meeting CDC guidelines for moderate- or vigorous-intensity aerobic activity, derived from the International Physical Activity Questionnaire (IPAQ). Percentages may not sum to 100% due to rounding.

Socioeconomic indicators varied across race-gender groups, with white men having the highest proportion of participants reporting no problem meeting financial needs (21.5%) and among Black men the highest proportion reporting frequent problems meeting financial needs (23.4%). White men also had the highest average educational attainment with 43.0% reporting a graduate degree, while Black men had the highest proportion of less than or equal to a high school education (22.3%). Higher perceived neighborhood problems were reported by white men (22.1%) and Black men (15.5%), while White women most frequently reported no perceived neighborhood problems (37.6%).

Health behaviors and reported health conditions also varied by race-gender group. Current smoking was most prevalent among White men (38.0%) and Black men (35.9%), while meeting physical activity guidelines was most common among White men (72.4%) and least common among Black women (57.2%). Fair or poor health was more commonly reported by women (22.8% of Black women and 26.5% of White women compared with 12.8% and 14.3% of Black and White men, respectively). The prevalence of diabetes was highest among white men (31.4%) and Black women (15.2%). Hypertension was most prevalent among Black women (32.4%) and least prevalent in Black men (23.7%).

### Main Effects

In main effects models, the association between perceived neighborhood problems (PNP) and hypertension varied substantially by race-gender (**Table 2**). Among White women and White men, greater PNP was consistently associated with increased odds of hypertension. In fully adjusted models, White women showed elevated odds at all levels: low PNP (aOR=2.31; 95% CI:1.29-4.13), moderate PNP (aOR=2.05 (1.17; 3.62)), and high PNP (aOR=2.88; 95% CI: 1.44, 5.74), compared with no PNP. White men similarly demonstrated increased odds of hypertension across all PNP levels. In contrast, among Black women and men, associations between PNP and hypertension were inconsistent. Black women showed lower odds at moderate PNP in unadjusted models (OR=0.57; 95% CI: 0.35, 0.94), but the effect estimate attenuated after adjustment.

**Table 2.**
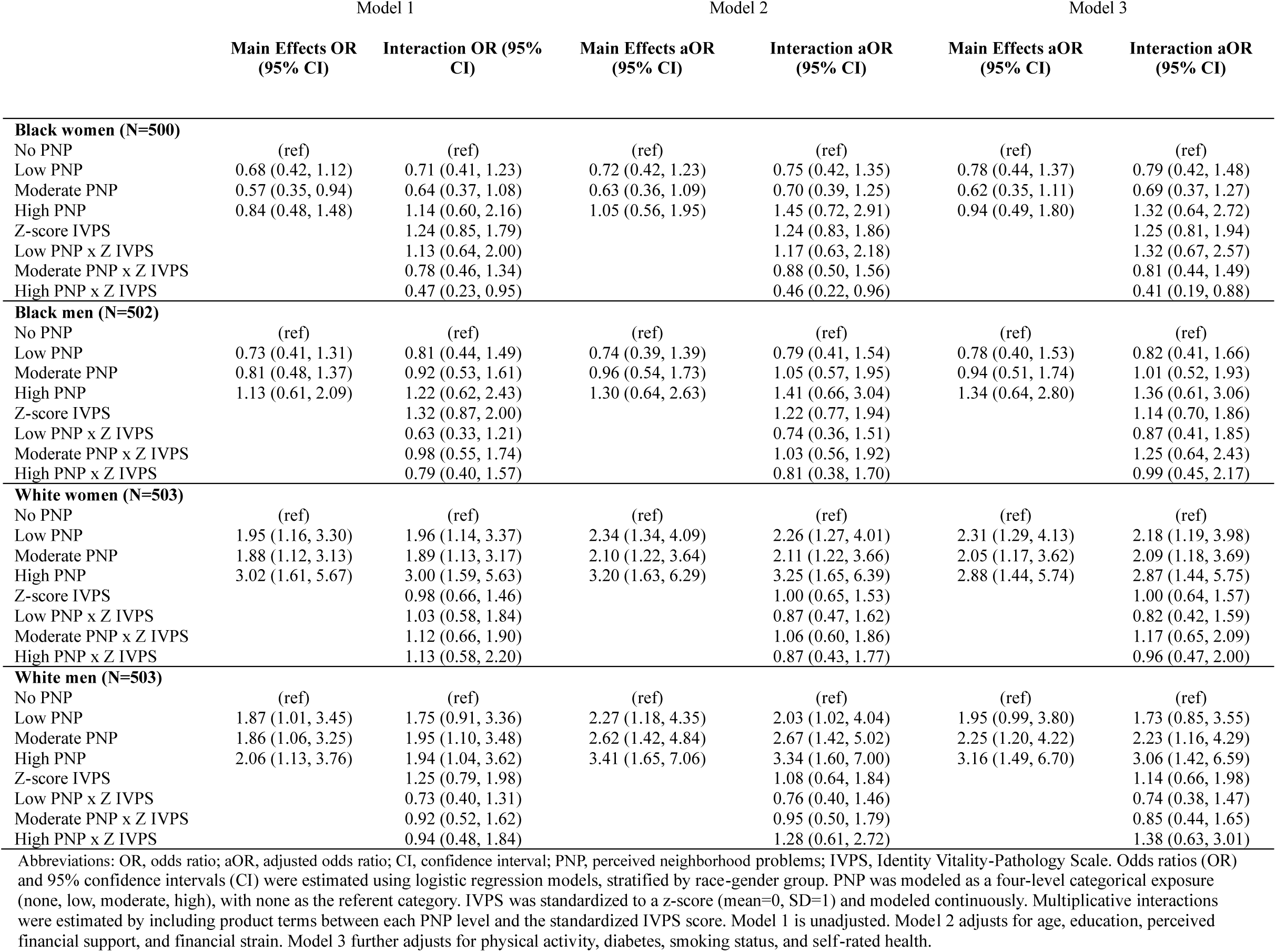
Association Between Perceived Neighborhood Problems and Hypertension, and Moderation by Identity State, Stratified by Race-Gender Group.

|  | Model 1 |  | Model 2 |  | Model 3 |  |
| --- | --- | --- | --- | --- | --- | --- |
|  | Main Effects OR<br>(95% CI) | Interaction OR (95%<br>CI) | Main Effects aOR<br>(95% CI) | Interaction aOR<br>(95% CI) | Main Effects aOR<br>(95% CI) | Interaction aOR<br>(95% CI) |
| **Black women (N=500)** |  |  |  |  |  |  |
| No PNP | (ref) | (ref) | (ref) | (ref) | (ref) | (ref) |
| Low PNP | 0.68 (0.42, 1.12) | 0.71 (0.41, 1.23) | 0.72 (0.42, 1.23) | 0.75 (0.42, 1.35) | 0.78 (0.44, 1.37) | 0.79 (0.42, 1.48) |
| Moderate PNP | 0.57 (0.35, 0.94) | 0.64 (0.37, 1.08) | 0.63 (0.36, 1.09) | 0.70 (0.39, 1.25) | 0.62 (0.35, 1.11) | 0.69 (0.37, 1.27) |
| High PNP | 0.84 (0.48, 1.48) | 1.14 (0.60, 2.16) | 1.05 (0.56, 1.95) | 1.45 (0.72, 2.91) | 0.94 (0.49, 1.80) | 1.32 (0.64, 2.72) |
| Z-score IVPS |  | 1.24 (0.85, 1.79) |  | 1.24 (0.83, 1.86) |  | 1.25 (0.81, 1.94) |
| Low PNP x Z IVPS |  | 1.13 (0.64, 2.00) |  | 1.17 (0.63, 2.18) |  | 1.32 (0.67, 2.57) |
| Moderate PNP x Z IVPS |  | 0.78 (0.46, 1.34) |  | 0.88 (0.50, 1.56) |  | 0.81 (0.44, 1.49) |
| High PNP x Z IVPS |  | 0.47 (0.23, 0.95) |  | 0.46 (0.22, 0.96) |  | 0.41 (0.19, 0.88) |
| **Black men (N=502)** |  |  |  |  |  |  |
| No PNP | (ref) | (ref) | (ref) | (ref) | (ref) | (ref) |
| Low PNP | 0.73 (0.41, 1.31) | 0.81 (0.44, 1.49) | 0.74 (0.39, 1.39) | 0.79 (0.41, 1.54) | 0.78 (0.40, 1.53) | 0.82 (0.41, 1.66) |
| Moderate PNP | 0.81 (0.48, 1.37) | 0.92 (0.53, 1.61) | 0.96 (0.54, 1.73) | 1.05 (0.57, 1.95) | 0.94 (0.51, 1.74) | 1.01 (0.52, 1.93) |
| High PNP | 1.13 (0.61, 2.09) | 1.22 (0.62, 2.43) | 1.30 (0.64, 2.63) | 1.41 (0.66, 3.04) | 1.34 (0.64, 2.80) | 1.36 (0.61, 3.06) |
| Z-score IVPS |  | 1.32 (0.87, 2.00) |  | 1.22 (0.77, 1.94) |  | 1.14 (0.70, 1.86) |
| Low PNP x Z IVPS |  | 0.63 (0.33, 1.21) |  | 0.74 (0.36, 1.51) |  | 0.87 (0.41, 1.85) |
| Moderate PNP x Z IVPS |  | 0.98 (0.55, 1.74) |  | 1.03 (0.56, 1.92) |  | 1.25 (0.64, 2.43) |
| High PNP x Z IVPS |  | 0.79 (0.40, 1.57) |  | 0.81 (0.38, 1.70) |  | 0.99 (0.45, 2.17) |
| **White women (N=503)** |  |  |  |  |  |  |
| No PNP | (ref) | (ref) | (ref) | (ref) | (ref) | (ref) |
| Low PNP | 1.95 (1.16, 3.30) | 1.96 (1.14, 3.37) | 2.34 (1.34, 4.09) | 2.26 (1.27, 4.01) | 2.31 (1.29, 4.13) | 2.18 (1.19, 3.98) |
| Moderate PNP | 1.88 (1.12, 3.13) | 1.89 (1.13, 3.17) | 2.10 (1.22, 3.64) | 2.11 (1.22, 3.66) | 2.05 (1.17, 3.62) | 2.09 (1.18, 3.69) |
| High PNP | 3.02 (1.61, 5.67) | 3.00 (1.59, 5.63) | 3.20 (1.63, 6.29) | 3.25 (1.65, 6.39) | 2.88 (1.44, 5.74) | 2.87 (1.44, 5.75) |
| Z-score IVPS |  | 0.98 (0.66, 1.46) |  | 1.00 (0.65, 1.53) |  | 1.00 (0.64, 1.57) |
| Low PNP x Z IVPS |  | 1.03 (0.58, 1.84) |  | 0.87 (0.47, 1.62) |  | 0.82 (0.42, 1.59) |
| Moderate PNP x Z IVPS |  | 1.12 (0.66, 1.90) |  | 1.06 (0.60, 1.86) |  | 1.17 (0.65, 2.09) |
| High PNP x Z IVPS |  | 1.13 (0.58, 2.20) |  | 0.87 (0.43, 1.77) |  | 0.96 (0.47, 2.00) |
| **White men (N=503)** |  |  |  |  |  |  |
| No PNP | (ref) | (ref) | (ref) | (ref) | (ref) | (ref) |
| Low PNP | 1.87 (1.01, 3.45) | 1.75 (0.91, 3.36) | 2.27 (1.18, 4.35) | 2.03 (1.02, 4.04) | 1.95 (0.99, 3.80) | 1.73 (0.85, 3.55) |
| Moderate PNP | 1.86 (1.06, 3.25) | 1.95 (1.10, 3.48) | 2.62 (1.42, 4.84) | 2.67 (1.42, 5.02) | 2.25 (1.20, 4.22) | 2.23 (1.16, 4.29) |
| High PNP | 2.06 (1.13, 3.76) | 1.94 (1.04, 3.62) | 3.41 (1.65, 7.06) | 3.34 (1.60, 7.00) | 3.16 (1.49, 6.70) | 3.06 (1.42, 6.59) |
| Z-score IVPS |  | 1.25 (0.79, 1.98) |  | 1.08 (0.64, 1.84) |  | 1.14 (0.66, 1.98) |
| Low PNP x Z IVPS |  | 0.73 (0.40, 1.31) |  | 0.76 (0.40, 1.46) |  | 0.74 (0.38, 1.47) |
| Moderate PNP x Z IVPS |  | 0.92 (0.52, 1.62) |  | 0.95 (0.50, 1.79) |  | 0.85 (0.44, 1.65) |
| High PNP x Z IVPS |  | 0.94 (0.48, 1.84) |  | 1.28 (0.61, 2.72) |  | 1.38 (0.63, 3.01) |

Abbreviations: OR, odds ratio; aOR, adjusted odds ratio; CI, confidence interval; PNP, perceived neighborhood problems; IVPS, Identity Vitality-Pathology Scale. Odds ratios (OR) and 95% confidence intervals (CI) were estimated using logistic regression models, stratified by race-gender group. PNP was modeled as a four-level categorical exposure (none, low, moderate, high), with none as the referent category. IVPS was standardized to a z-score (mean=0, SD=1) and modeled continuously. Multiplicative interactions were estimated by including product terms between each PNP level and the standardized IVPS score. Model 1 is unadjusted. Model 2 adjusts for age, education, perceived financial support, and financial strain. Model 3 further adjusts for physical activity, diabetes, smoking status, and self-rated health.

### Moderation by identity state

The inclusion of identity state as a moderator revealed that the relationship between PNP and hypertension differed meaningfully based on IVPS score, but only among Black women (**Table 2**). Within this group, identity state modified the relationship between PNP and hypertension (interaction OR for high vs. no PNP = 0.47, 95% CI: 0.23, 0.95), with the magnitude of the association magnified in fully adjusted models (interaction aOR=0.41; 95% CI 0.19, 0.88). This interaction indicates that among Black women perceiving the highest level of neighborhood problems, for each 1-SD increase in IVPS score, there was 59% lower odds of hypertension. Figure 2 illustrates this association with an aOR of 0.23 (95% CI: 0.06, 0.93) among Black women with a more vitalized identity state (+2 SD above the mean) and an aOR of 7.68 (95% CI: 1.13, 52.16) among Black women with a more pathologized identity state (–2 SD below the mean). In contrast, no meaningful modification of the PNP-hypertension association by identity state was observed among Black men, White women, or White men. Notably, there is a similar pattern of modification among Black men and White women (**Figure 2**).

**Fig. 2.**
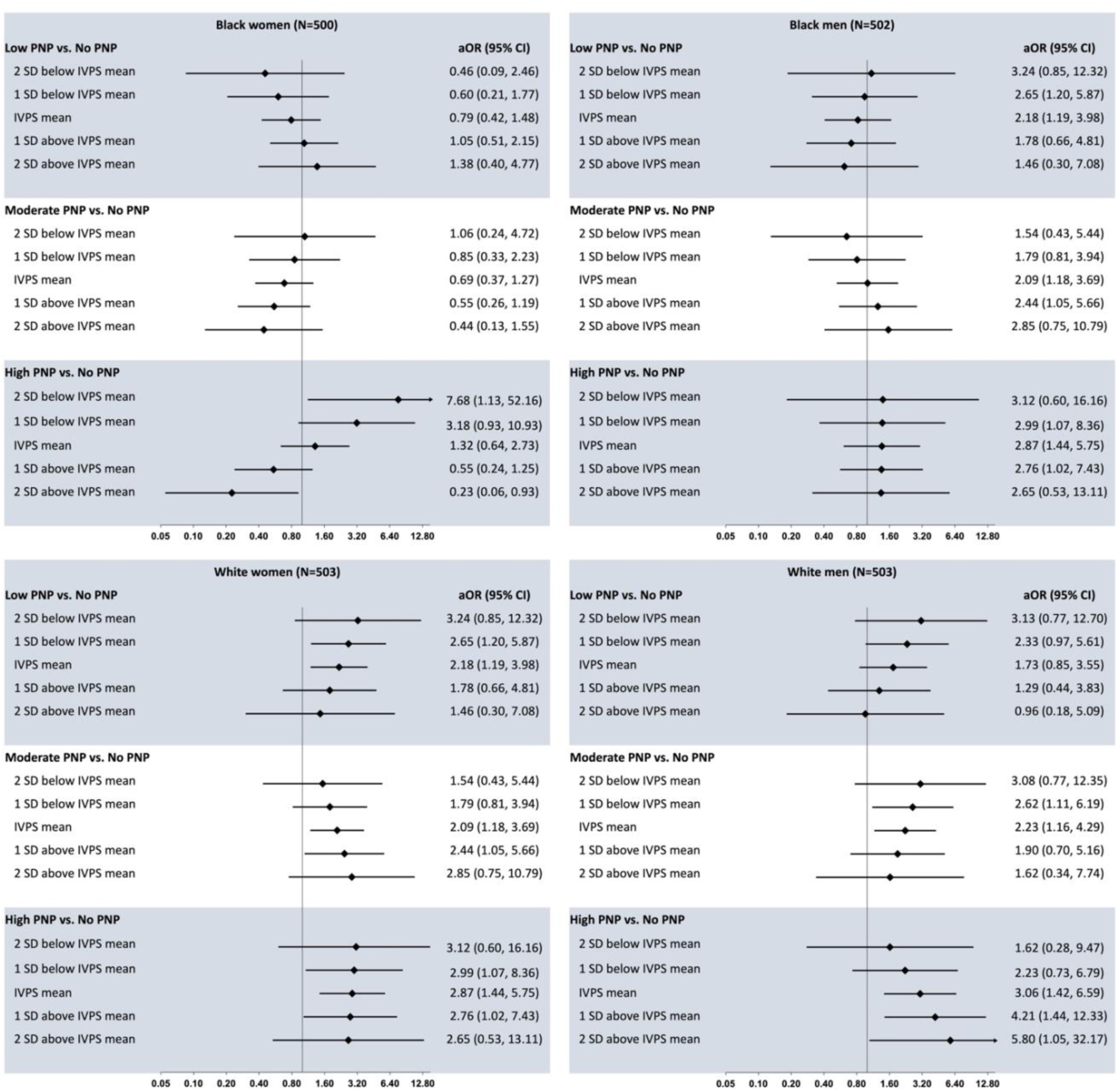
Adjusted Odds Ratios and 95% Confidence Intervals for Perceived Neighborhood Problems by Identity Vitality Pathology Score, Stratified by Race-Gender Group. Abbreviations: PNP, perceived neighborhood problems; SD, standard deviation; IVPS, identity vitality pathology score. PNP was modeled as a four-level categorical exposure (none, low, moderate, high), with none as the referent category. IVPS was standardized to a z-score (mean=0, SD=1) and modeled continuously. Multiplicative interactions were estimated by including product terms between each PNP level and the standardized IVPS score. Adjusts for age, education, perceived financial support, financial strain, physical activity, diabetes, smoking status, and self-rated health.

## Discussion

In this study, we examined the association between subjective experiences of neighborhood disadvantage, measured using the PNP scale, and hypertension across race-gender groups and evaluated whether identity state, as proposed by the Identity Vitality Pathology framework [37], moderated these associations. Given that the PNP captures the experiential dimensions of neighborhood disadvantage and that physiological stress responses are initiated through cognitive appraisal, we hypothesized that identity vitality would attenuate the PNP-hypertension association by modifying stress perception upstream of neuroendocrine activation. Consistent with prior literature linking neighborhood context to cardiovascular outcomes [16,47,48], we found that higher perceived neighborhood problems were associated with greater odds of hypertension, though this association varied across race-gender groups. Further, consistent with our hypothesis, we found race-gender differences in the degree to which a vitalized identity state is associated with decreased odds of hypertension in the context of perceived neighborhood disadvantage.

Among White women and men, the association between high PNP and hypertension was large, positive, and robust. In contrast, we found no evidence of an association between perceiving a high level of neighborhood problems and hypertension across all three levels of adjustment among Black women and men. With stress as a key risk factor for hypertension [49], effective intervention requires consideration of why perceiving more neighborhood problems may be more stress-promoting among White persons than Black persons. Previous literature suggests that greater neighborhood disorder is associated with feeling less safe among non-Hispanic (NH) White persons, but not NH Black persons [50]. Further exploration of the mechanisms underpinning these ethnoracial differences found that neighborhood socioeconomic status is not associated with safety perceptions when holding racial composition and crime rates constant [51]. This phenomenon is likely informed by the conflation of race and criminality perpetuated by mass media [52] and misconceived causal conclusions drawn from an association between neighborhood crime rates and racial composition that is, in reality, heavily confounded by racialized disinvestment [51]. Notably, White men were most likely to perceive neighborhood problems while White women were least likely, suggesting that subjective experiences of neighborhood adversity are highly gendered, which is an existing finding in the literature [53,54] that warrants further investigation.

Differences in how neighborhood threats are perceived and appraised in the context of chronic exposure to structural and community-level adversity may provide another potential explanation for the lack of association between perceived neighborhood problems and hypertension we observed among Black persons. This possibility is supported by prior research documenting attenuated perceptions of threat in response to violent crime among Black residents of Black neighborhoods [51] as well as lower-than-expected levels of depressive symptoms among Black adolescents exposed to the highest levels of community violence [55]. These findings suggest that repeated exposure to neighborhood adversity may shape how threats are perceived and appraised, potentially contributing to differences in the relationship between perceived neighborhood problems and health across racial groups.

However, these racial patterns warrant closer examination in the context of the observed moderation by identity state, where a consistent pattern emerged across all models. The positive association between subjective experiences of neighborhood disadvantage and hypertension among Black women and men may have been obscured by the omission of relevant factors that influence the relationship, such as group-specific impact of resilience resources. At mean levels of the IVPS score, high perceived neighborhood problems were positively associated with hypertension in the fully adjusted moderation model, suggesting that an underlying positive association may exist but was obscured in the main effects models which did not account for potential moderation by resilience factors such as identity state. The models examining moderation by identity state, in contrast, indicate that among Black women who perceive high levels of neighborhood problems, a more vitalized identity state was associated with a markedly lower prevelance of hypertension. This finding, showing that a highly vitalized identity state was most protective against hypertension at the highest levels of perceived neighborhood disadvantage, is consistent with the IVP framework’s proposition that identity vitality mitigates physiological stress reactivity at the greatest levels of stress exposure and buffers the unique manifestations of stress within a specific intersectional ethnoracial-gender group [36,37]. These findings suggest that the lack of an observed association between subjective neighborhood disadvantage and hypertension among Black women in this study reflects not an absence of neighborhood-related hypertension risk, but rather the presence of a psychosocial resilience resource that meaningfully and uniquely impacts the cardiometabolic health of Black women at the highest levels of perceived neighborhood disadvantage. Importantly, identity state appears to operate differently on the cardiometabolic health of other race-gender groups, who may manifest their stress as different conditions (e.g. premature heart failure among Black men [56]). This hypothesis is supported by our finding that the inclusion of identity state in models among Black men, White men, and White women did not alter main effect estimates, underscoring the necessity of considering heterogeneity across intersectional identities when examining psychosocial resilience. Notably, the pattern of moderation observed among Black men closely paralleled that of White women, a finding also consistent with the IVP framework’s assertion that these two groups may share underlying identity pathologies rooted in their simultaneous occupation of dominant and subordinate social positions [37]. This shared vulnerability, stemming from an idealized but inaccessible social status, may help explain why identity state operates similarly to shape cardiometabolic risk across these disparate sociodemographic groups.

Other resilience resources have been identified in relation to hypertension outcomes among Black Americans. Specifically, interpersonal social support has been found to buffer the relationship between socioeconomic disadvantage and elevated blood pressure [57], while neighborhood organizational participation has similarly been linked to lower hypertension risk [35]. However, both of these resources depend on the presence of external relational or civic infrastructure, which are patterned by the same neighborhood disinvestment driving the health disparities in question [58]. Conversely, identity vitality enables access to stress mitigation whether or not external resources are available. A vitalized identity state reduces the likelihood that a stressor tied to one’s social position (i.e. neighborhood disadvantage) is perceived as threatening, widens an individual’s sense of control and self-efficacy when a threat is perceived, and, in turn, allows for a broader range of adaptive coping responses [36]. Because identitiy state is theorized as a modifiable characteristic that functions intrinsically, promoting identity vitality may represent a more directly intervenable target that can be implemented across neighborhood environments where health-determining material resources are least available. This upstream, identity-based positioning makes identity state moderation particularly relevant to understanding resilience to the physiological manifestations of stress among Black women in the context of neighborhood disadvantage.

Black women occupy a unique position at the intersection of racial and gendered oppression, and the cumulative weight of these systems has long shaped both the health burdens carried [59] and, consequentially, the necessitated coping resources [60]. These systems of oppression do not operate independently; rather, they combine into a form of gendered racism that is distinct from either racism or sexism alone, which works through direct and indirect pathways to erode health and well-being [61]. Racial and gender discrimination increases vulnerability to stressors across multiple domains of daily life, including financial hardship, social network loss, and victimization [62], suggesting that structural disadvantage does not stay confined to one domain, but instead proliferates into the everyday circumstances of Black women’s lives over time. For example, Black women have been disproportionately targeted by predatory lending practices [63] and face higher rates of eviction [64] than any of the other race-gender groups in our study, meaning the housing instability produced by structural racism is not experienced uniformly across Black communities, but falls with particular force on Black women. It is precisely within this unique context that psychosocial resources rooted in identity may matter most. Black women navigating the chronic stress of multiply disadvantaged social positions may stand to benefit most from a vitalized identity state that shapes how structurally-rooted stressors are perceived, potentially interrupting the pathways from chronic stress stemming from neighborhood disadvantage to prevalent adverse physiological outcomes such as hypertension.

Several strengths and limitations of this study warrant consideration. A notable strength of this analysis is the use of the HARI dataset, which intentionally oversampled Black participants, providing the statistical power necessary to examine heterogeneity in the association of subjective neighborhood disadvantage with hypertension across race-gender groups. Hypertension status was ascertained by self-report, which is known to have imperfect sensitivity [65]. Racial disparities in undiagnosed hypertension have largely been attributed to differences in socioeconomic position rather than race itself, with prior work findings that racial differences in hypertension rates are attenuated substantially once neighborhood affluence and other socioenvironmental factors are accounted for [66]. Given that the HARI study sample was constructed to balance socioeconomic status across race-gender groups, differential misclassification of hypertention status by race or gender is unlikely to be a major source of bias, though some degree of nondifferential misclassification cannot be ruled out.

While not intended to be nationally representative, HARI participants were drawn from all four U.S. census regions, which allows for reasonably broad extrapolation of our findings to Black and White adults across the U.S. with internet access. Because the study oversampled Black women and men and low-SES White women and men, we were able to more precisely assess how race, gender, and SES intersect in patterning hypertension risk, given the more comparable covariate distributions this produced across groups. This quota sampling method provided the power to detect subgroup differences, but also introduced selection bias, and applying weighting techniques to generalize to the broader U.S. population would be misleading given this design.[67] Online recruitment alsp carries potential for selection bias, namely differential access to and engagement with research panels [68]. While weighting could inadequately address this concern, the limited generalizability of online surveys is well established and we consider a more appropriate approach to interpret our results as reflective of digitally literate individuals with internet access, rather than the U.S. population at large [68].

Given our study design, we also do not anticpate differential selection bias based on online recruitment across the race-gender groups. Panel access and participation tend to track more with socioeconomic factors like education and internet availability, as well as sociocultural factors like English fluency rather than race or gender [69,70]. Eligibility criteria of the HARI study included non-Hispanic ethnicity and English fluency, therefore language was unlikely to differentially shape recruitment across our race-gender groups. Our recruitment partner also used demographic profiling, stratified sampling, and geographically targeted recruitment to reach harder-to-reach populations [71], and we intentionally recruited comparable socioeconomic profiles across the four subgroups. Together, these strategies reduce the likelihood of differential selection bias by race and gender.

This study provides evidence that subjective experiences of neighborhood disadvantage are heterogeneously associated with hypertension, and that identity vitality, a novel, modifiable identity-based resilience phenotype, attenuates this association among Black women specifically. We interpret our finding that this moderation was not observed among other race-gender groups as theoretically consistent rather than incidental, reflecting the unique position Black women maintain at the cross-section of structural disadvantage and social stressors that may make psychosocial resources rooted in identity particularly beneficial for cardiometabolic health. Future work should prioritize longitudinal designs that can establish the temporal ordering of neighborhood socioeconomic status, identity state, and hypertension onset, and should consider formally evaluating both moderating and mediating roles of psychosocial resilience resources in the pathway from structural disadvantage to biological risk. Ultimately, these findings point toward the possibility that interventions aimed at cultivating identity vitality among Black women living in disinvested neighborhoods may represent a meaningful, if partial, path for reducing cardiovascular health disparities rooted in structural racism.

## Data Availability

All data produced in the present study are available upon reasonable request to the authors and will be made available upon the completion of ongoing analyses

## Acknowledgements

The Healthy Aging with Resilient Identities Study and author G.B. were supported by National Institute on Aging contract R00AG075327. Author H.P. is supported by a National Heart, Lung, and Blood Institute T32 grant (HL007055). The authors thank the participants of the HARI study for their valuable contributions.

## Author contributions

All authors contributed to the study conception and design, with the primary design led by Ganga Bey. Data analysis was performed by Hannah Pleasants. The first draft of the manuscript was written by Hannah Pleasants, and Ganga Bey commented and revised previous versions of the manuscript. All authors read and approved the final manuscript.

## References

1. Martin SS, Aday AW, Allen NB, Almarzooq ZI, Anderson CAM, Arora P, et al. 2025 Heart Disease and Stroke Statistics: A Report of US and Global Data From the American Heart Association. Circulation. 2025 Feb 25;151(8):e41–660. doi:10.1161/CIR.0000000000001303 PubMed PMID: 39866113; PubMed Central PMCID: PMC12256702.

2. Hardy ST, Jaeger BC, Foti K, Ghazi L, Wozniak G, Muntner P. Trends in Blood Pressure Control among US Adults With Hypertension, 2013-2014 to 2021-2023. Am J Hypertens. 2025 Jan 16;38(2):120–8. doi:10.1093/ajh/hpae141 PubMed PMID: 39504487; PubMed Central PMCID: PMC11735471.

3. Noah GU, Omohoro MU, Magacha HM, Fuko CD, Ezike T. Racial Disparities in Hypertension-Related Hospital Mortality Among Adults in the United States. Cureus. 17(3):e81043. doi:10.7759/cureus.81043 PubMed PMID: 40264630; PubMed Central PMCID: PMC12014224.

4. Mukaz DK, Sparks AD, Plante TB, Judd SE, Howard G, Howard VJ, et al. Residential Racial Segregation, Socioeconomic Status, and Hypertension Risk in Black and White Americans: The REGARDS Prospective Cohort Study. J Am Heart Assoc. 2025 Oct 7;14(19):e041339. doi:10.1161/JAHA.125.041339

5. Smith GS, McCleary RR, Thorpe RJ. Racial Disparities in Hypertension Prevalence within US Gentrifying Neighborhoods. Int J Environ Res Public Health. 2020 Nov;17(21):7889. doi:10.3390/ijerph17217889 PubMed PMID: 33126467; PubMed Central PMCID: PMC7662342.

6. Rothstein R. The Color of Law: A Forgotten History of How Our Government Segregated America. Liveright Publishing; 2017. 243 p.

7. Lawrence T., Brown P. The Black Butterfly [Internet]. Johns Hopkins University Press; 2022 [cited 2026 Feb 18]. Available from: https://www.press.jhu.edu/books/title/11971/black-butterfly doi:10.56021/9781421439884

8. University of California Press [Internet]. [cited 2026 Apr 23]. Golden Gulag by Ruth Gilmore – Paper. Available from: https://www.ucpress.edu/books/golden-gulag/paper

9. Riley T, Schleimer JP, Jahn JL. Organized abandonment under racial capitalism: Measuring accountable actors of structural racism for public health research and action. Soc Sci Med. 2024 Feb 1;343:116576. doi:10.1016/j.socscimed.2024.116576

10. Odoms-Young A, Brown AGM, Agurs-Collins T, Glanz K. Food Insecurity, Neighborhood Food Environment, and Health Disparities: State of the Science, Research Gaps and Opportunities. Am J Clin Nutr. 2024 Mar 1;119(3):850–61. doi:10.1016/j.ajcnut.2023.12.019

11. Kirby JB, Kaneda T. Neighborhood socioeconomic disadvantage and access to health care. J Health Soc Behav. 2005 Mar;46(1):15–31. doi:10.1177/002214650504600103 PubMed PMID: 15869118.

12. Turrell G, Haynes M, Burton NW, Giles-Corti B, Oldenburg B, Wilson LA, et al. Neighborhood disadvantage and physical activity: baseline results from the HABITAT multilevel longitudinal study. Ann Epidemiol. 2010 Mar;20(3):171–81. doi:10.1016/j.annepidem.2009.11.004 PubMed PMID: 20159488.

13. Lee JGL, Henriksen L, Rose SW, Moreland-Russell S, Ribisl KM. A Systematic Review of Neighborhood Disparities in Point-of-Sale Tobacco Marketing. Am J Public Health. 2015 Sep;105(9):e8–18. doi:10.2105/AJPH.2015.302777 PubMed PMID: 26180986; PubMed Central PMCID: PMC4529779.

14. Alaniz ML. Alcohol Availability and Targeted Advertising in Racial/Ethnic Minority Communities. Alcohol Health Res World. 1998;22(4):286–9. PubMed PMID: 15706757; PubMed Central PMCID: PMC6761895.

15. Wheeler DC, Boyle J, Barsell DJ, Glasgow T, McClernon FJ, Oliver JA, et al. Associations of Alcohol and Tobacco Retail Outlet Rates with Neighborhood Disadvantage. Int J Environ Res Public Health. 2022 Jan 20;19(3):1134. doi:10.3390/ijerph19031134 PubMed PMID: 35162162; PubMed Central PMCID: PMC8834944.

16. Mujahid MS, Diez Roux AV, Morenoff JD, Raghunathan TE, Cooper RS, Ni H, et al. Neighborhood characteristics and hypertension. Epidemiology. 2008 Jul;19(4):590–8. doi:10.1097/EDE.0b013e3181772cb2 PubMed PMID: 18480733.

17. Diez Roux AV, Mair C. Neighborhoods and health. Ann N Y Acad Sci. 2010;1186(1):125–45. doi:10.1111/j.1749-6632.2009.05333.x

18. Weber J, Angerer P, Apolinário-Hagen J. Physiological reactions to acute stressors and subjective stress during daily life: A systematic review on ecological momentary assessment (EMA) studies. PLoS ONE. 2022 Jul 27;17(7):e0271996. doi:10.1371/journal.pone.0271996 PubMed PMID: 35895674; PubMed Central PMCID: PMC9328558.

19. Andrews MR, Ceasar J, Tamura K, Langerman SD, Mitchell VM, Collins BS, et al. Neighborhood environment perceptions associate with depression levels and cardiovascular risk among middle-aged and older adults: Data from the Washington, DC cardiovascular health and needs assessment. Aging Ment Health. 2021 Nov;25(11):2078–89. doi:10.1080/13607863.2020.1793898 PubMed PMID: 32691611; PubMed Central PMCID: PMC7855489.

20. McEwen BS. Allostasis and Allostatic Load: Implications for Neuropsychopharmacology. Neuropsychopharmacology. 2000 Feb;22(2):108–24. doi:10.1016/S0893-133X(99)00129-3

21. Inoue K, Horwich T, Bhatnagar R, Bhatt K, Goldwater D, Seeman T, et al. Urinary Stress Hormones, Hypertension, and Cardiovascular Events: The Multi-Ethnic Study of Atherosclerosis. Hypertension. 2021 Nov;78(5):1640–7. doi:10.1161/HYPERTENSIONAHA.121.17618

22. McEwen BS. Stress, adaptation, and disease. Allostasis and allostatic load. Ann N Y Acad Sci. 1998 May 1;840:33–44. doi:10.1111/j.1749-6632.1998.tb09546.x PubMed PMID: 9629234.

23. Mocayar Marón FJ, Ferder L, Saraví FD, Manucha W. Hypertension linked to allostatic load: from psychosocial stress to inflammation and mitochondrial dysfunction. Stress. 2019 Mar;22(2):169–81. doi:10.1080/10253890.2018.1542683 PubMed PMID: 30547701.

24. Dallman MF, Akana SF, Laugero KD, Gomez F, Manalo S, Bell ME, et al. A spoonful of sugar: feedback signals of energy stores and corticosterone regulate responses to chronic stress. Physiol Behav. 2003 Jun 1; Proceedings from the 2002 Meeting of the Society for the Study of Ingestive Behavior (SSIB)79(1):3–12. doi:10.1016/S0031-9384(03)00100-8

25. Dallman MF, Pecoraro N, Akana SF, la Fleur SE, Gomez F, Houshyar H, et al. Chronic stress and obesity: A new view of “comfort food.” Proc Natl Acad Sci U S A. 2003 Sep 30;100(20):11696–701. doi:10.1073/pnas.1934666100 PubMed PMID: 12975524; PubMed Central PMCID: PMC208820.

26. Jackson JS, Knight KM, Rafferty JA. Race and Unhealthy Behaviors: Chronic Stress, the HPA Axis, and Physical and Mental Health Disparities Over the Life Course. Am J Public Health. 2010 May;100(5):933–9. doi:10.2105/AJPH.2008.143446 PubMed PMID: 19846689; PubMed Central PMCID: PMC2853611.

27. Forde AT, Sims M, Muntner P, Lewis T, Onwuka A, Moore K, et al. Discrimination and Hypertension Risk among African Americans in the Jackson Heart Study. Hypertens Dallas Tex 1979. 2020 Sep;76(3):715–23. doi:10.1161/HYPERTENSIONAHA.119.14492 PubMed PMID: 32605388; PubMed Central PMCID: PMC8359680.

28. Social Determinants of Risk and Outcomes for Cardiovascular Disease. Circulation [Internet]. [cited 2026 Aug 4]. Available from: https://www.ahajournals.org/doi/10.1161/CIR.0000000000000228

29. Kershaw KN, Robinson WR, Gordon-Larsen P, Hicken MT, Goff DC, Carnethon MR, et al. Association of Changes in Neighborhood-Level Racial Residential Segregation With Changes in Blood Pressure Among Black Adults: The CARDIA Study. JAMA Intern Med. 2017 Jul 1;177(7):996–1002. doi:10.1001/jamainternmed.2017.1226 PubMed PMID: 28505341; PubMed Central PMCID: PMC5710452.

30. Sims M, Diez-Roux AV, Gebreab SY, Brenner A, Dubbert P, Wyatt S, et al. Perceived Discrimination is Associated with Health Behaviors among African Americans in the Jackson Heart Study. J Epidemiol Community Health. 2016 Feb;70(2):187–94. doi:10.1136/jech-2015-206390 PubMed PMID: 26417003; PubMed Central PMCID: PMC5014355.

31. Hernandez R, González HM, Tarraf W, Moskowitz JT, Carnethon MR, Gallo LC, et al. Association of dispositional optimism with Life’s Simple 7’s Cardiovascular Health Index: results from the Hispanic Community Health Study/Study of Latinos (HCHS/SOL) Sociocultural Ancillary Study (SCAS). BMJ Open. 2018 Mar 5;8(3):e019434. doi:10.1136/bmjopen-2017-019434 PubMed PMID: 29567845.

32. Rozanski A, Bavishi C, Kubzansky LD, Cohen R. Association of Optimism With Cardiovascular Events and All-Cause Mortality: A Systematic Review and Meta-analysis. JAMA Netw Open. 2019 Sep 27;2(9):e1912200. doi:10.1001/jamanetworkopen.2019.12200 PubMed PMID: 31560385.

33. Pleasants H, Pike JR, Palta P, Bertoni AG, Hughes TM, Xiao Q, et al. Psychosocial Risk and Resilience as Moderators of the Association Between Neighborhood Disadvantage and Incident Cardiovascular Disease Across Ethnoracial Groups: Multi-Ethnic Study of Atherosclerosis, United States, 2000-2019. Am J Public Health. 2026 May;116(5):711–21. doi:10.2105/AJPH.2025.308407 PubMed PMID: 41950447; PubMed Central PMCID: PMC13066695.

34. Coulon SM, Wilson DK, Alia KA, Van Horn ML. Multilevel Associations of Neighborhood Poverty, Crime, and Satisfaction With Blood Pressure in African-American Adults. Am J Hypertens. 2016 Jan;29(1):90–5. doi:10.1093/ajh/hpv060 PubMed PMID: 25917562; PubMed Central PMCID: PMC5014129.

35. Sharp G, Carpiano RM. Neighborhood social organization exposures and racial/ethnic disparities in hypertension risk in Los Angeles. PLOS ONE. 2023 Mar 6;18(3):e0282648. doi:10.1371/journal.pone.0282648 PubMed PMID: 36877695; PubMed Central PMCID: PMC9987829.

36. Bey G, Rivadeneira N, Berman C, Carmody M, Brondolo E. Initial Validation of the Long-form and Short-form Identity Vitality-Pathology Scale [Internet]. 2025 Oct 19 [cited 2026 Aug 4]. Available from: https://osf.io/gmup5_v1

37. Bey GS. The Identity Vitality-Pathology model: A novel theoretical framework proposing “identity state” as a modulator of the pathways from structural to health inequity. Soc Sci Med 1982. 2022 Nov 1;314:115495. doi:10.1016/j.socscimed.2022.115495 PubMed PMID: 36335704.

38. Crenshaw K. Demarginalizing the Intersection of Race and Sex: A Black Feminist Critique of Antidiscrimination Doctrine, Feminist Theory and Antiracist Politics.

39. Woods-Giscombé CL. Superwoman Schema: African American Women’s Views on Stress, Strength, and Health. Qual Health Res. 2010 May;20(5):668–83. doi:10.1177/1049732310361892 PubMed PMID: 20154298; PubMed Central PMCID: PMC3072704.

40. Woods-Giscombe CL, Williams KP, Conklin J, Dodd A, Bravo L, Anderson AM, et al. A scoping review of the concept of resilience among African American women. Arch Psychiatr Nurs. 2023 Oct;46:107–20. doi:10.1016/j.apnu.2023.04.008 PubMed PMID: 37813493; PubMed Central PMCID: PMC11587904.

41. Farag NH, Moore WE, Lovallo WR, Mills PJ, Khandrika S, Eichner JE. Hypothalamic-Pituitary-Adrenal Axis Function: Relative Contributions of Perceived Stress and Obesity in Women. J Womens Health. 2008 Dec;17(10):1647–55. doi:10.1089/jwh.2008.0866 PubMed PMID: 19049359; PubMed Central PMCID: PMC2945932.

42. Martin CL, Ghastine L, Wegienka G, Wise LA, Baird DD, Vines AI. Early Life Disadvantage and the Risk of Depressive Symptoms among Young Black Women. J Racial Ethn Health Disparities. 2024 Jun 1;11(3):1819–28. doi:10.1007/s40615-023-01654-x

43. Xiao Y (Karen), Graham G. Where we live: The impact of neighborhoods and community factors on cardiovascular health in the United States. Clin Cardiol. 2018 Nov 29;42(1):184–9. doi:10.1002/clc.23107 PubMed PMID: 30393880; PubMed Central PMCID: PMC6436513.

44. 44. Disorder and Decay – Catherine E. Ross, John Mirowsky, 1999 [Internet]. [cited 2026 Feb 24]. Available from: https://journals.sagepub.com/doi/10.1177/107808749903400304

45. Craig CL, Marshall AL, Sjöström M, Bauman AE, Booth ML, Ainsworth BE, et al. International Physical Activity Questionnaire: 12-Country Reliability and Validity. Med Sci Sports Exerc. 2003 Aug;35(8):1381. doi:10.1249/01.MSS.0000078924.61453.FB

46. Physical Activity Guidelines for Americans, 2nd edition.

47. Kim Y, Twardzik E, Judd SE, Colabianchi N. Neighborhood Socioeconomic Status and Stroke Incidence. Neurology. 2021 May 11;96(19):897–907. doi:10.1212/WNL.0000000000011892 PubMed PMID: 33766995; PubMed Central PMCID: PMC8166445.

48. Coulon SM, Wilson DK, Alia KA, Van Horn ML. Multilevel Associations of Neighborhood Poverty, Crime, and Satisfaction With Blood Pressure in African-American Adults. Am J Hypertens. 2016 Jan;29(1):90–5. doi:10.1093/ajh/hpv060 PubMed PMID: 25917562; PubMed Central PMCID: PMC5014129.

49. Marwaha K. Examining the Role of Psychosocial Stressors in Hypertension. J Prev Med Pub Health. 2022 Nov 30;55(6):499–505. doi:10.3961/jpmph.21.266

50. Velasquez AJ, Douglas JA, Guo F, Robinette JW. In the eyes of the beholder: Race, place and health. Front Public Health. 2022 Aug 12;10:920637. doi:10.3389/fpubh.2022.920637 PubMed PMID: 36033798; PubMed Central PMCID: PMC9412158.

51. Cho W, Ho AT. Does neighborhood crime matter? A multi-year survey study on perceptions of race, victimization, and public safety. Int J Law Crime Justice. 2018 Dec 1;55:13–26. doi:10.1016/j.ijlcj.2018.08.002

52. Weiss A, Chermak SM. The News Value of African-American Victims: An Examination of the Media’s Presentation of Homicide. J Crime Justice. 1998 Jan 1;21(2):71–88. doi:10.1080/0735648X.1998.9721601

53. Jackson AL, Soller B, Browning CR. The Influence of Women’s Neighborhood Resources on Perceptions of Social Disorder. City Community. 2017 Jun;16(2):189–208. doi:10.1111/cico.12229 PubMed PMID: 28757810; PubMed Central PMCID: PMC5528866.

54. Stafford M, Cummins S, Macintyre S, Ellaway A, Marmot M. Gender differences in the associations between health and neighbourhood environment. Soc Sci Med. 2005 Apr 1;60(8):1681–92. doi:10.1016/j.socscimed.2004.08.028

55. Gaylord-Harden NK, Dickson D, Pierre C. Profiles of Community Violence Exposure Among African American Youth: An Examination of Desensitization to Violence Using Latent Class Analysis. J Interpers Violence. 2016 Jul 1;31(11):2077–101. doi:10.1177/0886260515572474

56. Understanding the Complexity of Heart Failure Risk and Treatment in Black Patients. Circ Heart Fail [Internet]. [cited 2026 Sep 9]. Available from: https://www.ahajournals.org/doi/10.1161/CIRCHEARTFAILURE.120.007264

57. Byrd DR. Social Support Satisfaction as a Buffer Against Hypertension-Related Cognitive Decline in Black Americans. Alzheimers Dement. 2025 Dec 23;21(Suppl 6):e107454. doi:10.1002/alz70860_107454 PubMed PMID: null; PubMed Central PMCID: PMC12726149.

58. Hailu EM, Reeves AN, McAlexander T, Judd S, Odden MC. Neighborhood Disinvestment and Racial Disparities in Early Hypertension Onset Among Women. JAMA Netw Open. 2026 Jun 23;9(6):e2619845. doi:10.1001/jamanetworkopen.2026.19845

59. Geronimus AT. The weathering hypothesis and the health of African-American women and infants: evidence and speculations. Ethn Dis. 1992;2(3):207–21. PubMed PMID: 1467758.

60. Woods-Giscombe CL, Williams KP, Conklin J, Dodd A, Bravo L, Anderson AM, et al. A scoping review of the concept of resilience among African American women. Arch Psychiatr Nurs. 2023 Oct;46:107–20. doi:10.1016/j.apnu.2023.04.008 PubMed PMID: 37813493; PubMed Central PMCID: PMC11587904.

61. Chinn JJ, Martin IK, Redmond N. Health Equity Among Black Women in the United States. J Womens Health. 2021 Feb 1;30(2):212–9. doi:10.1089/jwh.2020.8868 PubMed PMID: 33237831; PubMed Central PMCID: PMC8020496.

62. Perry BL, Harp KLH, Oser CB. Racial and Gender Discrimination in the Stress Process: Implications for African American Women’s Health and Well-Being. Sociol Perspect SP Off Publ Pac Sociol Assoc. 2013;56(1):25–48. PubMed PMID: 24077024; PubMed Central PMCID: PMC3783344.

63. Phillips S. The Subprime Mortgage Calamity and the African American Woman. Rev Black Polit Econ. 2012 Jan 1;39(2):227–37. doi:10.1007/s12114-011-9107-1

64. Hepburn P, Louis R, Desmond M. Racial and Gender Disparities among Evicted Americans. Sociol Sci. 2020 Dec 16;7:649–62. doi:10.15195/v7.a27

65. Vargas CM, Burt VL, Gillum RF, Pamuk ER. Validity of Self-Reported Hypertension in the National Health and Nutrition Examination Survey III, 1988–1991. Prev Med. 1997 Sep 1;26(5):678–85. doi:10.1006/pmed.1997.0190

66. Jeong S, Linder BA, Barnett AM, Tharpe MA, Hutchison ZJ, Culver MN, et al. Interplay of Race and Neighborhood Deprivation on Ambulatory Blood Pressure in Young Adults. medRxiv. 2023 Sep 12;2023.09.11.23295160. doi:10.1101/2023.09.11.23295160 PubMed PMID: 37745604; PubMed Central PMCID: PMC10516077.

67. Baker R, Brick JM, Bates NA, Battaglia M, Couper MP, Dever JA, et al. Summary Report of the AAPOR Task Force on Non-probability Sampling. J Surv Stat Methodol. 2013 Nov 1;1(2):90–143. doi:10.1093/jssam/smt008

68. Bethlehem J. Selection Bias in Web Surveys. Int Stat Rev. 2010;78(2):161–88. doi:10.1111/j.1751-5823.2010.00112.x

69. Prepared for the AAPOR Executive Council by a Task Force operating under the auspices of the AAPOR Standards Committee, with members including:, Baker R, Blumberg SJ, Brick JM, Couper MP, Courtright M, et al. Research Synthesis: AAPOR Report on Online Panels. Public Opin Q. 2010 Dec 1;74(4):711–81. doi:10.1093/poq/nfq048

70. Hays RD, Liu H, Kapteyn A. Use of Internet panels to conduct surveys. Behav Res Methods. 2015 Sep;47(3):685–90. doi:10.3758/s13428-015-0617-9 PubMed PMID: 26170052; PubMed Central PMCID: PMC4546874.

71. ESOMAR. Konovo [Internet]. [cited 2026 Aug 4]. Available from: https://konovo.com/esomar/

